# Impact and Assessment of Mechanical Tension in Bowel Anastomosis: a Scoping Review

**DOI:** 10.1101/2024.06.12.24308856

**Authors:** Muhammad Usman Khalid, Danish Ali, Jie Ying Wu, Aimal Khan

## Abstract

**Background:** While tension on anastomoses is primarily regarded as a critical factor in anastomotic leaks and failure, assessing this tension is based on subjective surgeon estimation. There is currently no clinically available tool to assess mechanical tension on an anastomosis objectively. Some animal and human studies have previously evaluated anastomotic tension, but no comprehensive review discusses the different methods and types of tension measured.

**Objectives:** To summarize the current state of the literature regarding the measurement and impact of tension on bowel anastomoses.

**Design:** This scoping review was conducted with a systematic search of literature in the PubMed, SCOPUS, and EMBASE databases. Key terms used were anastomoses/anastomosis, tension, bowel, surgery, intestine, tissue, small bowel, large bowel, bowel, mucosal tissue, and other corollary terms. Data were synthesized in tables, and summarized paragraphically, with studies assessed using the New-Castle Ottawa scale where possible. Emphasis was placed on method of tension assessment, outcomes, and factors relating to tension.

**Results:** Anastomotic leak and tension are strongly associated, with the presence of tension making leaks up to 10 times more likely. While freedom from tension has traditionally been measured via the surrogate measure of adequate bowel mobilization, this remains a subjective and imprecise method. The literature describes several techniques to allow adequate mobilization, such as splenic flexure mobilization or division of the omentum. However, basing the estimate of tension on subjective assessment has some inherent drawbacks. Animal and cadaveric studies have been the frontier for objective measurement of wall tension, with the use of scaffolds, suture types, and prostheses to bolster the natural tolerance of the bowel. However, these tend to use tensiometers to measure tension, along with automated machines or pulley and ratcheting systems to increase tension in specified intervals. These are universally destructive due to their design of measuring maximal tensile load as well as not being easily adaptable to the operating room.

**Conclusions:** The current literature does not study objective measurement of bowel tension in live human subjects. Bowel mobilization is a common method to reduce tension, but it relies on subjective judgment, which varies between surgeons. Given the recognized importance of tension, developing an objective, safe, intra-operative method to measure bowel wall tension would be a valuable surgical tool.

## Introduction

Anastomotic leak is a severe complication of gastrointestinal surgery, resulting in significant morbidity, mortality, and increased healthcare costs ^1-4^. Occurring in 2-30% of cases (depending on the location), anastomotic leaks increase the risk of mortality tenfold ^5-8^. Survivors face extended hospital stays, intensive care unit admissions, poor quality of life, and lasting disabilities ^4,9-11^. Additionally, leaks can lead to worse oncologic outcomes of reduced survival and higher recurrence rates ^12^. Each instance of leakage imposes a substantial economic burden, ranging from $25,000 to $150,000 per patient in increased hospital expenses. This amount does not consider the potential loss of productivity among survivors ^13-15^.

The defining principles behind successful anastomoses have traditionally involved patient factors and three vital technical elements: good technique, adequate blood supply, and avoidance of tension ^16-21^. Anastomotic leaks, therefore, stem from an interplay of patient-related and technical risk factors, including microcirculation, technique, and tension-free anastomosis ^22-24^. Despite significant advances in understanding and mitigating the patient-related risk factors for anastomotic leaks, the role of technical factors, particularly anastomotic tension, in the development of leaks remains poorly understood ^16,25-27^.

In recent years, significant advances have been made in developing new techniques for assessing blood supply measurement. However, an objective measurement of bowel measurement of bowel tension remains elusive ^28,29^. Mechanical tension on bowel anastomosis is among the strongest predictors of anastomotic complications, leading to a tenfold increase in the risk of leaks ^30-34^. Anastomotic tension is still measured mainly in the same way as in the last 150 years when Halstead laid out his principles of surgery through subjective surgeon estimation ^24,35,36^. This reliance on subjective parameters hinders scientific progress in verifiable assessment and understanding of the impact of tension on leaks ^37,38^. The existing *ex vivo* studies that measured tissue tension in bowel anastomosis tend to be destructive and animal-based ^35,39-43^.

An objective measurement of mechanical bowel wall tension intra-operatively is needed to allow for scientific data-driven recommendations in human subjects and enable surgeons to improve outcomes. In this scoping review, we summarized the current literature on assessing anastomotic tension and its impact on leaks to highlight the current gaps in research on this crucial topic.

## Methods

### Study design

We designed a scoping review to collate and assess the current state of the literature regarding the measurement and impact of tension on anastomotic outcomes. The study was designed according to the Preferred Reporting Items for Systematic Reviews and Meta-Analysis for Scoping Reviews (PRISMA-ScR) checklist, as denoted in Figure 1^44^.

**Figure 1.**
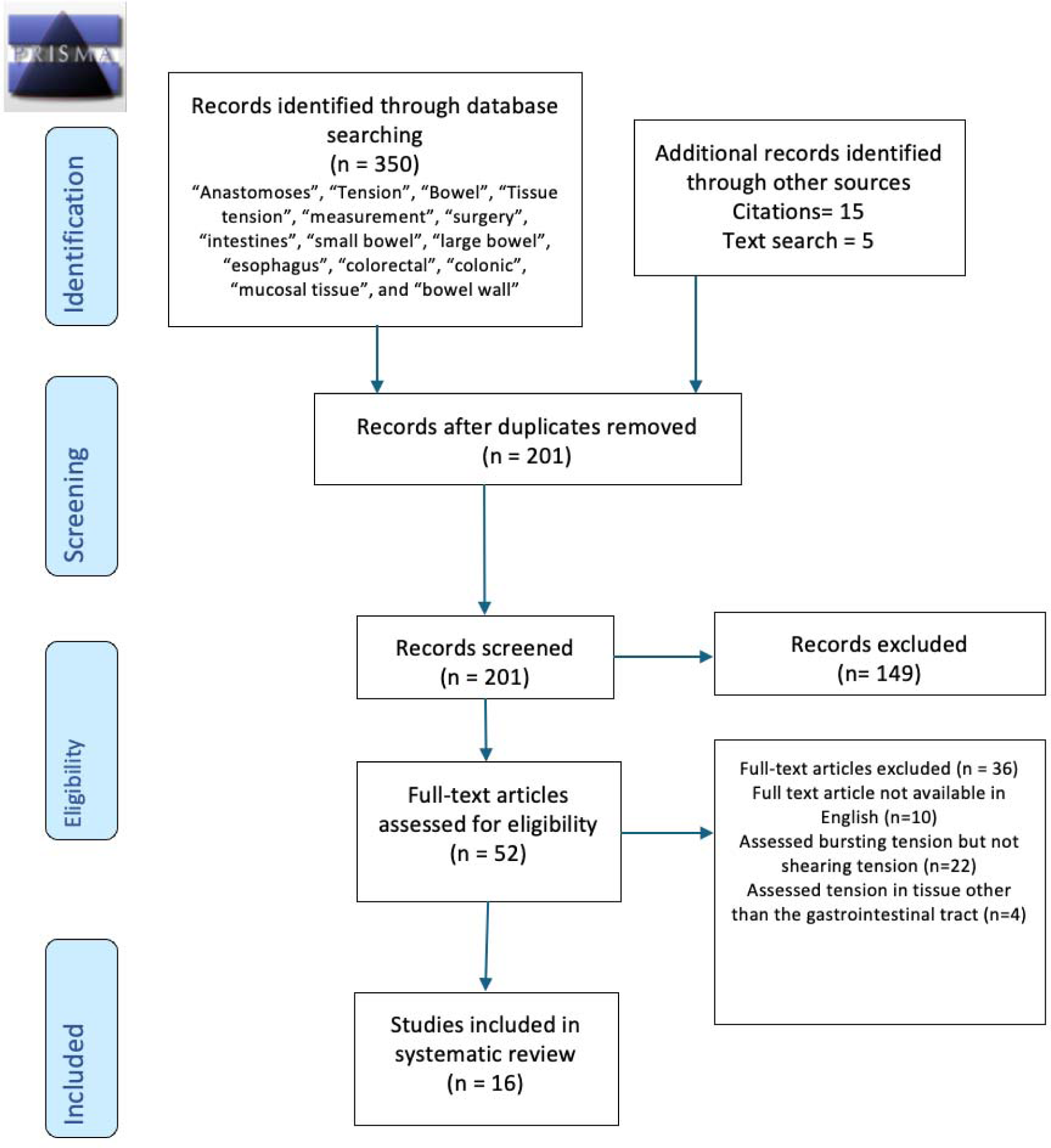
P Preferred Reporting Items for Systematic Reviews and Meta-Analysis for Scoping Reviews (PRISMA-ScR) flowchart

### Literature Search and Study Selection

An initial preliminary literature review was performed to identify subject-specific free-text and Medical Subject Headings (MeSH) terms. Subsequently, a bibliographic search was conducted across three electronic databases, MEDLINE (PubMed), SCOPUS, and EMBASE, using MeSH and free-text terms. All papers in the English language were considered, and there was no restriction on time period. Additionally, there were no restrictions on human, animal, in vivo, or ex vivo studies. References from published articles were also evaluated. Search terms included variations on the combinations of the terms (“anastomoses” OR “anastomosis”) AND “tension” AND “bowel” with optional terms using the operators AND/OR “tissue tension”, “measurement”, “surgery”, “intestine(s)”, “small bowel”, “large bowel”, “esophagus”, “colorectal”, “colonic”, “mucosal tissue”, and “bowel wall”.

Articles were collated and reviewed using the Rayyan.ai study screening tool. After removing 62 duplicates and articles not in English, a preliminary title review was performed by two authors DA and MK to eliminate irrelevant articles, excluding 107 articles. This was followed by an abstract review by the same authors, with 18 conflicts resolved by AK, in which 149 articles were removed as they did not mention anastomosis or tension. Finally, a full-text review was conducted on 52 studies based on relevance, with the results or discussion section mentioning anastomosis in the gastrointestinal tracts. Studies discussing bursting tension were excluded due to the inherent difference in the nature of the tension type. Furthermore, for inclusion, the article needed to discuss the presence or importance of anastomotic rupture, leak, or dehiscence. Selected articles were divided into two categories: studies examining the importance of tension in anastomosis in the bowel and studies with experimental techniques to investigate tension on anastomotic strength. Results were summarized in table 2.

### Quality Assessment

The quality of the studies included in our review was judged using the Newcastle Ottawa Scale (Table 1). Each study was judged on three broad perspectives: 1. Selection of the study groups, 2. Comparability of the groups, and 3. Ascertainment of the exposure or outcome of interest for case-control or cohort studies.

**Table 1:** Newcastle-Ottawa Scale for the studies included in this systematic review that met the study type qualifying criteria.

| Author | Representativeness of the exposed cohort | Selection of the nonexposed cohort | Ascertainment of exposure | Demonstration that Outcome of Interest was Not Present at the Start of Study | Comparability of Cohorts 1) Main Factor 2) Additional Factor | Assessment of Outcome | Sufficient Follow-Up | Adequacy of Follow-Up | Total 9/9 |
| --- | --- | --- | --- | --- | --- | --- | --- | --- | --- |
| Katory et al.<sup>48</sup> | * | * | * |  | * | * |  | * | 6/9 |
| Brennan et al.<sup>46</sup> | * |  | * |  |  | * | * | * | 5/9 |
| Oetzmann et al.<sup>52</sup> |  | * | * | * | ** | * |  |  | 5/9 |
| Egorov et al.<sup>70</sup> | * |  | * | * |  | * |  |  | 4/9 |
| Schilt et al.<sup>42</sup> | * | * | * | * | ** | * |  | * | 8/9 |
| Behrend et al.<sup>39</sup> |  | * | * | * | ** | * |  | * | 7/9 |
| Larsen et al.<sup>43</sup> |  |  | * | * | ** | * |  | * | 6/9 |
| Holland-Cunz et al.<sup>60</sup> |  | * | * | * | ** | * | * | * | 8/9 |
| Buch et al.<sup>40</sup> |  | * | * | * | ** | * |  | * | 7/9 |
| Waninger et al.<sup>41</sup> |  | * | * | * | ** | * |  |  | 6/9 |
| Morse et al.<sup>30</sup> | * |  | * |  | ** | * | * | * | 7/9 |

**Table 2:**
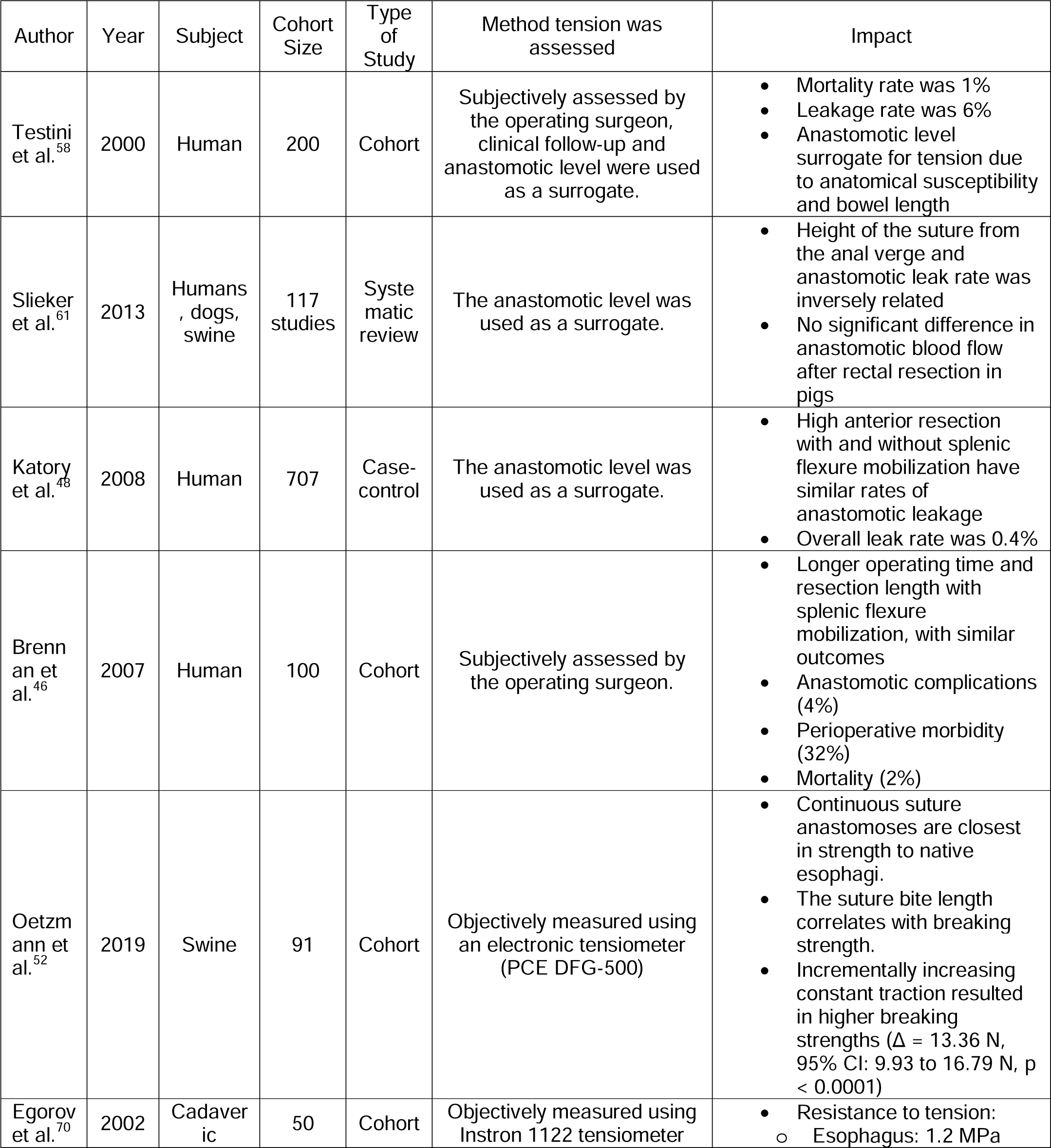
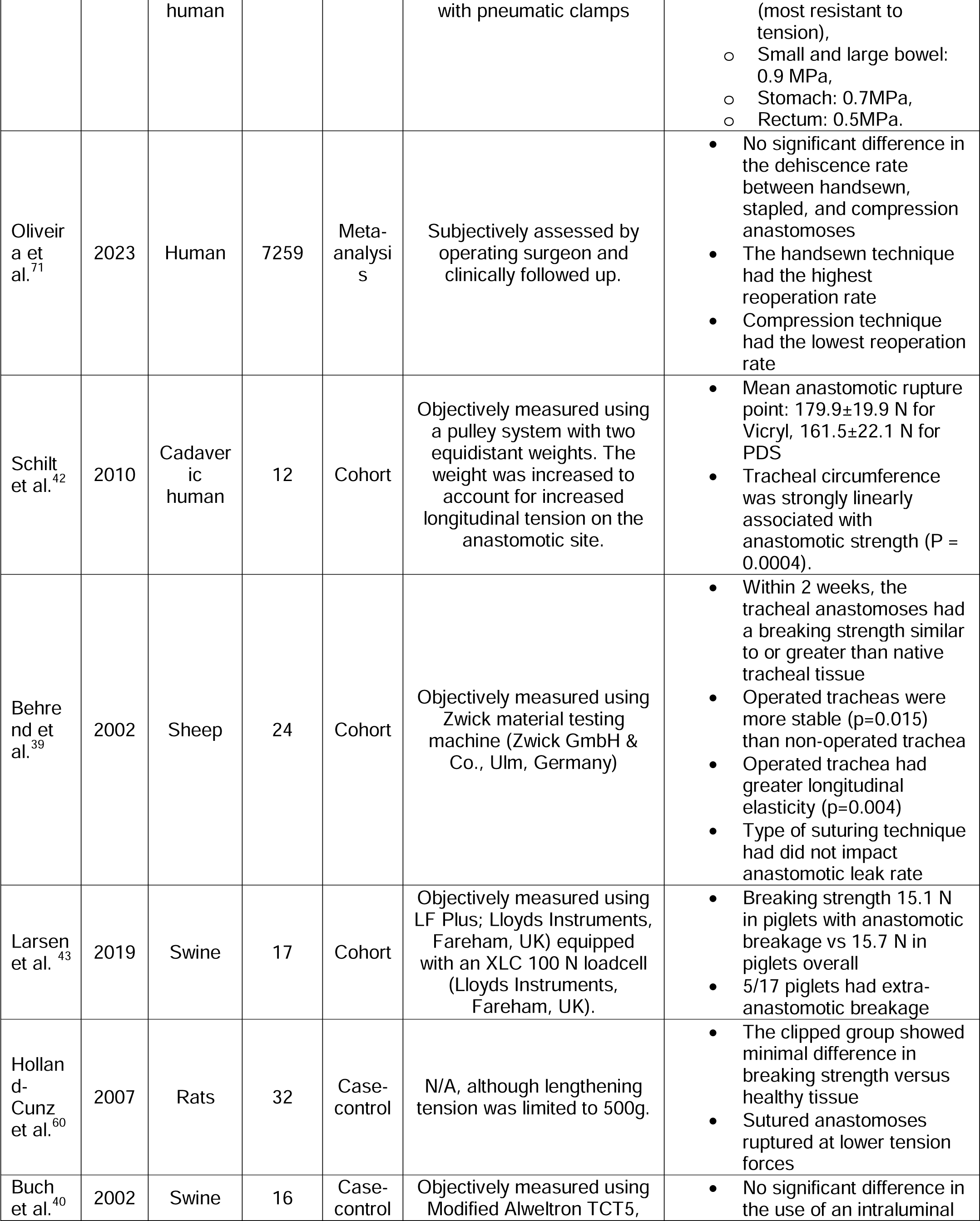
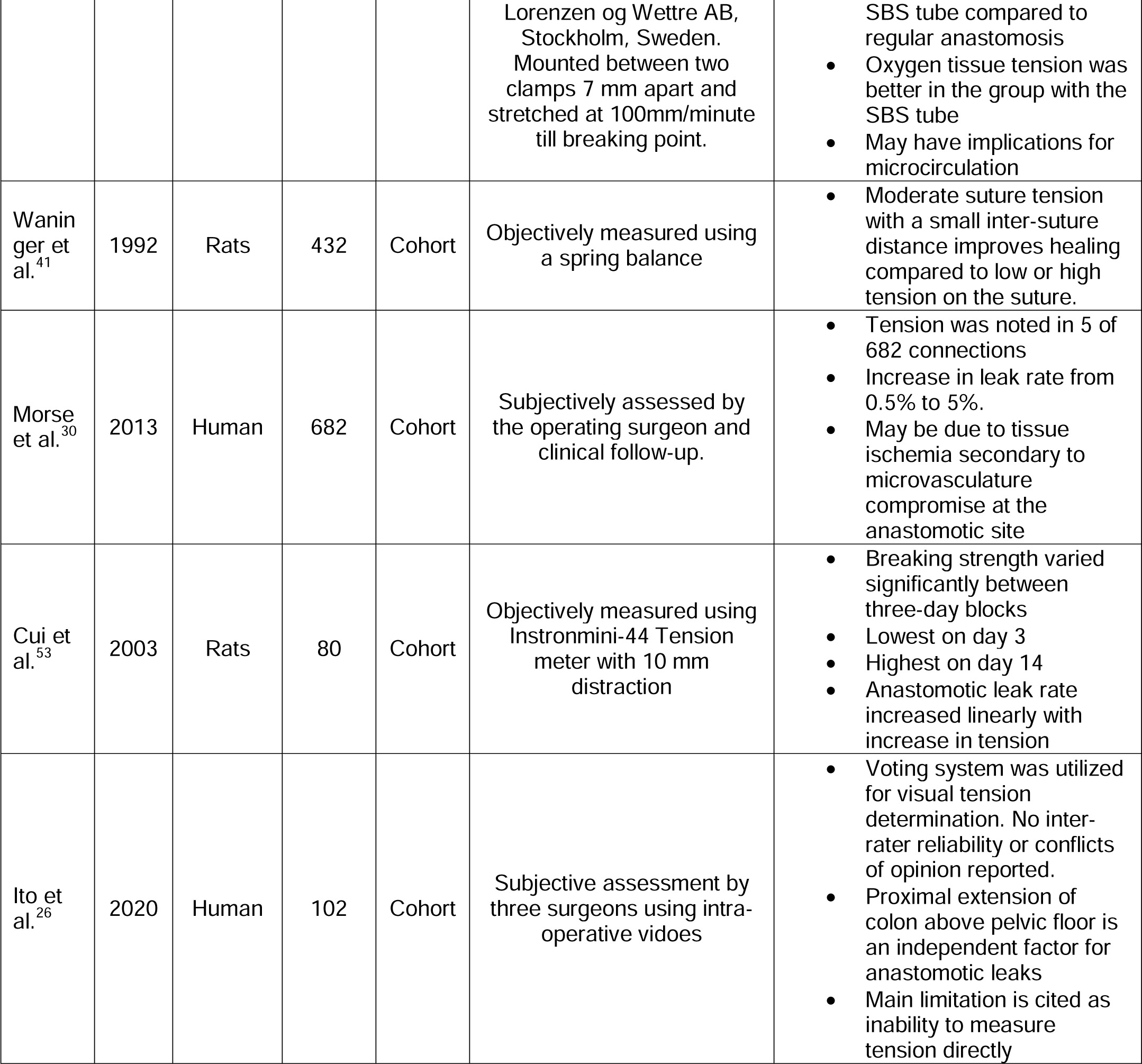
Summary of included studies, their year of publication, species studied, sample size, and significant findings.

### Findings

None of the studies conducted in humans to assess the impact of tension on bowel anastomosis measured tension directly or objectively. The method of tension measurement ranged from intra-operative subjective assessment to using surrogate markers like splenic flexure mobilization ^45,46^. The endpoint of tension assessment is usually a clinically apparent discharge from the anastomotic site in the form of gas, pus, or fecal matter, which irritates the peritoneal lining. A radiologic approach to detect a leak uses a hydrophilic compound on a Computed Tomography (CT) scan; however, it is only performed when a symptomatic leak is suspected ^8^.

Morse et al. found that the odds of intestinal anastomotic leakage were as much as 10.1 (CI: 1.3-76.9) with anastomotic tension, and the leak rate increased from 0.5% in patients with no tension on the anastomosis to 5% in patients with tension on the anastomosis (p=0.027) ^30^. Their study defined anastomotic leak by imaging, intra-operative extravasation of intra-luminal contents, and symptom-based findings. However, the study used a subjective assessment of anastomotic tension, based on imaging and surgeon assessment during reoperation for the leak, and had a low sample size of five patients with recorded anastomotic tension ^30^.

Wu et al. conducted a study to assess the impact of tension on clinical outcomes and quality of life among patients undergoing ileal pouch-anal anastomosis ^45^. Like Morse et al., they found that patients with high amounts of tension were more likely to suffer from anastomotic stricture and pouch failure. In the study, the tension on the bowel was an indirect measure of the surgeon’s assessment of mesenteric tension using a 10-point tension scale, with “1 being flappy and 10 being ‘as tight as a guitar string”.

Some studies have used mobilization of the colon as a surrogate for tension-free anastomoses. Splenic flexure mobilization is a routine part of anterior colorectal resection but is done at the expense of increased operating time, splenic injury, or even a splenectomy ^47^. A few authors seem to accept the trade-off of increased time to achieve adequate mobilization of surrounding structures to achieve a ‘tension-free’ anastomosis. However, they did not find a difference in complications with these measures ^48^.

Other surrogates for tension may be found in site selection. Low rectal anastomoses are more challenging to achieve tension-free and have higher rates of dehiscence compared to other rectal anastomoses ^49,50^. Generally, low rectal anastomoses are within 7 cm of the anal verge or below the peritoneal reflection ^51^. One way to counteract this issue is with a covering stoma, as indicated in many studies ^51^. Another region with higher tension anastomoses is the esophagus, having the highest failure rate of all gastrointestinal anastomoses ^4^. While other factors, such as delicate vasculature and technique, play a pivotal role in the anastomotic success in this region, the main distinction from other anastomoses is the higher anastomotic tension due to the anatomy, particularly post-operatively ^52^. The improved leak rate with load-bearing sutures to alleviate tension has corroborated this theory ^53,54^. Previous work with a tensiometer (to measure the tension) at different intervals and time points has demonstrated a linear increase of leak rates with tension on the anastomosis ^32,53^.

## Discussion

Tension, or lack thereof, in live human subjects is assessed intraoperatively by the surgeon. Factors that play a role in this assessment are anastomotic level, obesity, and anastomotic technique^7,10-12,16,55^. For patients with symptomatic anastomotic leaks post-operatively, the cause of tension may be analyzed. These studies tend to focus on demographic, operative, and disease related factors^19,46,48,56-59^. In animals anastomotic breaking strength is the subject investigated in literature, typically using machines to stretch bowel or tissue segments till rupture. This has been done in a variety of animals including rats, pigs, and dogs, generally reported in Newtons^22,33,39,41,43,52-54,60^. The common consensus is that tension plays a large role in anastomotic healing and leak rate, with higher tension causing adverse outcomes.

## The impact of tension on anastomotic leakage

Freedom from tension is achieved by adequate bowel mobilization, whether from the retroperitoneum, omentum, or other surrounding structures, based on the approach and type of surgery ^61^. Ileocolic anastomoses generally require mobilization of the colon of the retroperitoneum and possibly a division of the transverse colon omentum. The complexity increases for left-sided anastomoses where total mobilization of the descending colon and splenic flexure is required, leading to an increased risk of lower bowel perfusion. During a laparoscopic anterior resection, mobilization of the splenic flexure is often considered a vital step in achieving tension-free anastomoses. However, this maneuver increases operative time in exchange for a shorter length of stay; it has also not been shown to impact complications ^46,48,62^. These complex and time-consuming surgical techniques are intended to reduce tension but can significantly lengthen the procedure and carry risks ^63,64^. In a landmark study, Wu et al. investigated the correlation of the surgeon’s tension assessment with clinical outcomes in restorative proctocolectomy ^31^. Patients with high mesenteric tension were more likely to have anastomotic stricture and pouch failure, and unfavorable anatomy, such as a shorter anal transitional zone. Other patient factors, such as high body mass index, lower anastomosis level, longer distance between anastomotic segments, and lack of bowel mobilization, also independently increase the risk of complications ^17,34,57,59,65,66^. Conversely, surgical techniques that mitigate tension, such as increased bowel mobilization, splenic flexure mobilization, high arterial ligation, or tension-offloading sutures, yield reductions in leak rates ^25,67-69^. In all these approaches, the primary assessment is based on surgeon feedback, with the need for mobilization being an indirect method to alleviate or prevent tension. However, it is subjective and imprecise, based on surgical experience, comfort with the technique, and many other variables that can affect human judgement ^36^.

### Measurement of tension and impact on leaks

The most common objective way to measure tension in literature is using a tensiometer, a device with variability in its application and method. A standard procedure that is destructive and, therefore, commonly used posthumously is the attachment of the anatomy to a fixed and movable point. The displacement of the movable point exerting a counterforce is used to measure tension, as shown in Figure 2. This follows the assumption that the anastomosis is the weakest point of the anatomy and, therefore, the point of maximal tension; as such, the site of anastomotic leakage. However, only a few studies have evaluated these assumptions with material inside the anatomical segment ^53,54^. Additionally, most of these studies are focused on maximal tension bearing and rupture rather than a simple leak. Models of tensiometer commonly used include the Instron Mini 44 tensiometer (Canton, MA, USA) ^53^ and the Zwick material testing machine (Zwick GmbH & Co., Ulm, Germany) ^39^ A lower-cost model is visualized in Figure 3 and utilized for middle-lower income socioeconomic testing facilities. However, it comes at the cost of decreased sensitivity and more unaccounted variables. Mounting methods also differ, but are not as commonly described in the literature. Another variable is the advance rate of the movable portion of the machine, which could affect the tension rate. Some studies have found that where tension is increased in increments with pauses, the strength required to break the anastomosis is higher ^39,53^.

**Figure 2.**
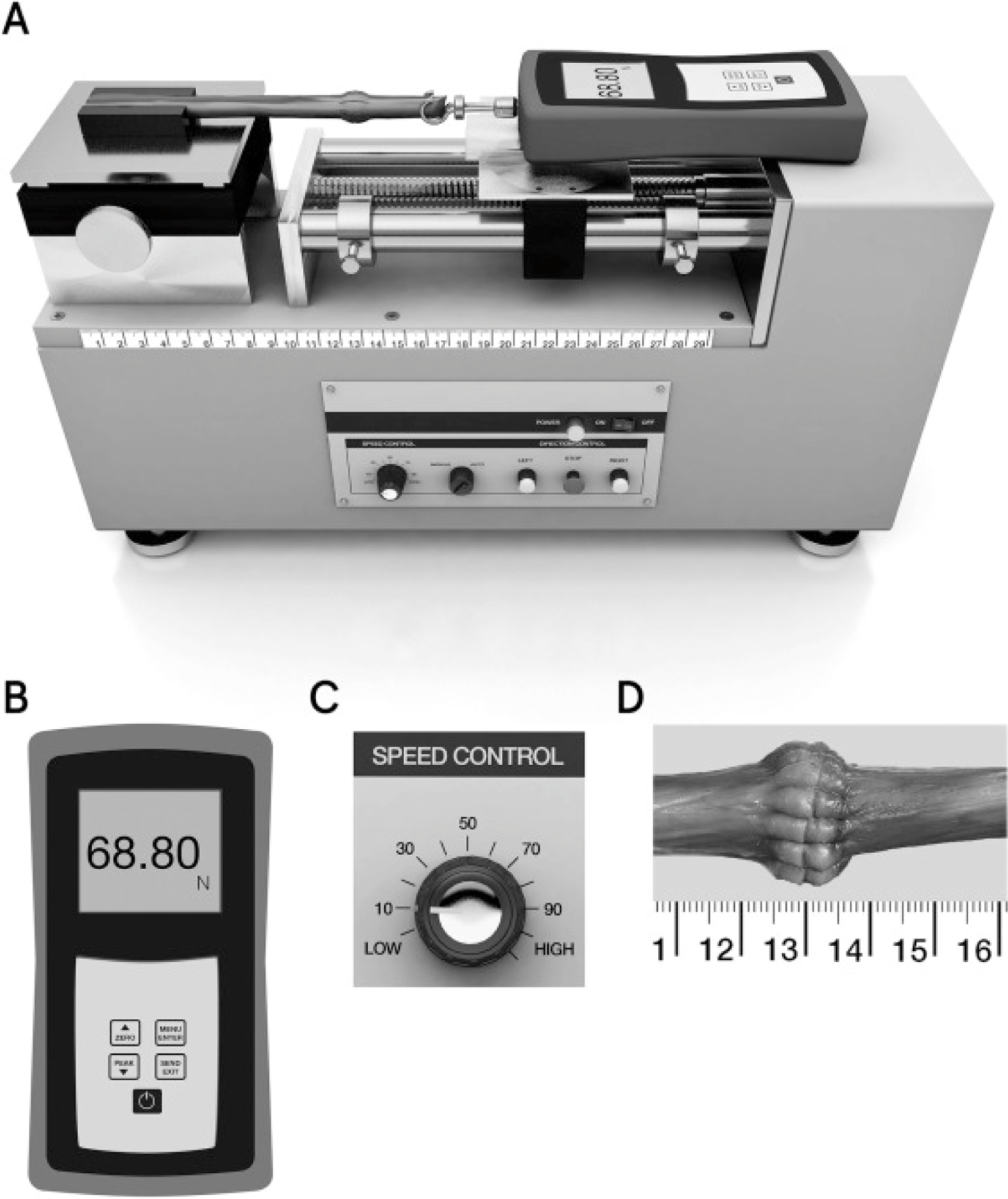
Electronic tensiometer with a horizontal clamping device presented by Oetzamann et al. Several key features are highlighted in Figures 2B, C, and D to reflect the tensiometer resolution, control over displacement speed, and anastomosis displacement measurement, respectively ^52^. *Reprinted with permission from Esophageal Biomechanics Revisited: A Tale of Tenacity, Anastomoses, and Suture Bite Lengths in Swine (1670-1677), by C. Oetzmann von Sochaczewski, 2019, The Annals of Thoracic Surgery. [2019] by Elsevier*.

**Figure 3.**
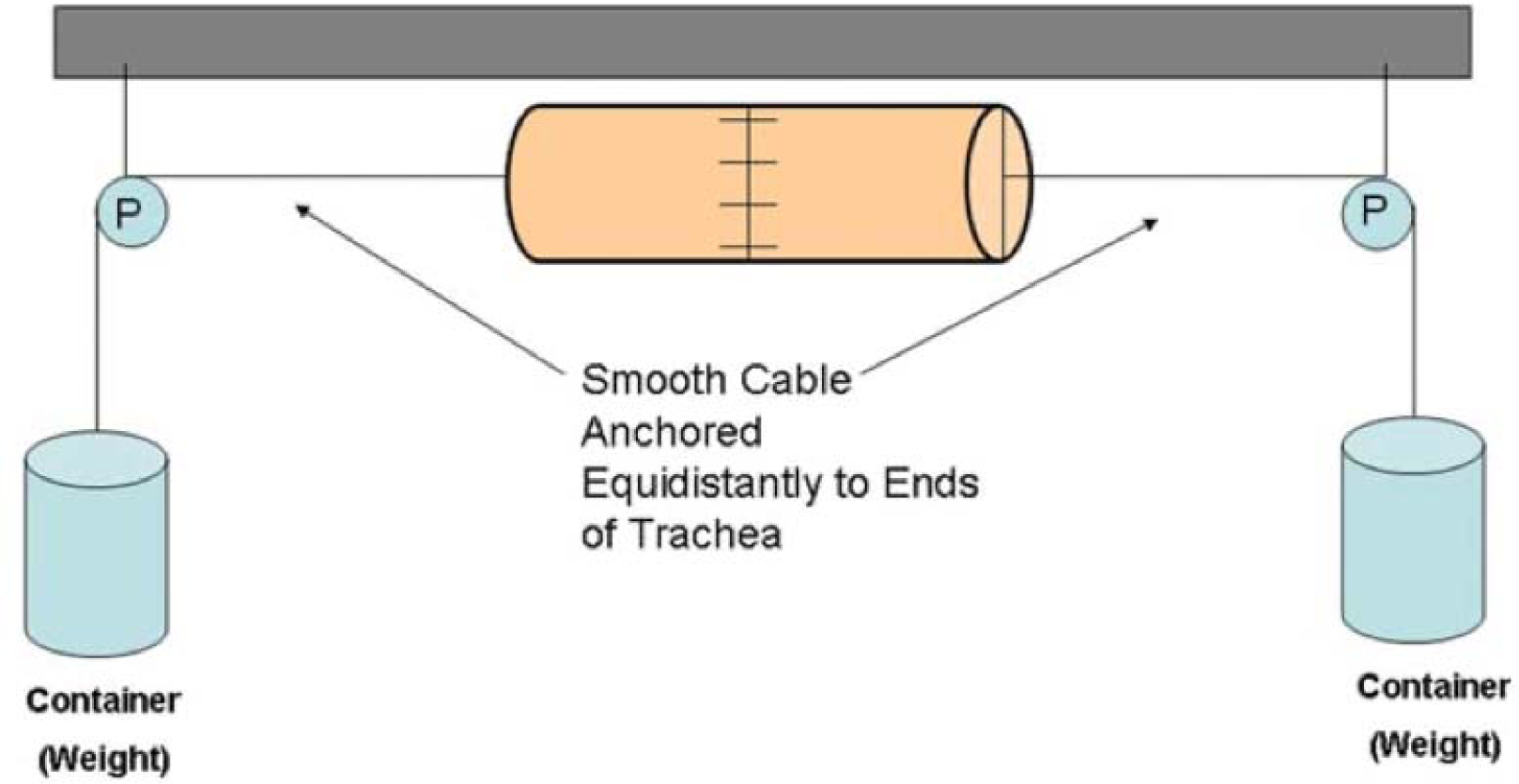
Low-cost pulley system as an alternative to tensiometers and mechanically operated displacement devices to increase and measure tension on the anastomosis presented by Schilt et al.^42^. *Reprinted with permission from An experimental model to investigate initial tracheal anastomosis strength (1125-1128), by P. N. Schilt, 2010, The Laryngoscope. [2010] by John Wiley and Sons*.

#### Suture tension and anastomotic type

Some articles correlate suture tension and variables such as suture length to investigate effects on anastomotic breaking strength. One rodent study investigated different suture tensions; moderate tension sutures gave optimal results ^41^. In the porcine esophagus, continuous sutures with short bites were closest to native tissue intolerance when investigating the maximum traction force tolerance in different suturing techniques and bite lengths ^52^. In this study, continuous sutures were significantly better than all other modalities – simple interrupted and barbed sutures or stapling ^52^. The esophagus, however, typically sustains higher traction forces than the stomach or bowel, which may skew the results ^70^. In other studies, similar postoperative complications have been noted in handsewn, stapled, and compression anastomotic techniques in meta-analyses ^71^. Another article compared the shearing forces of two common types of suture material, Vicryl (179.9 N) and Polydioxanone (161.5 N); the difference was not statistically significant ^42^. Most of the tissue tears were not at the site of the anastomoses but rather at a different site, which is concerning for the viability of the tissue tested ^42^. Another study compared a new clipped intestinal non-perforating technique but did not specify the testing parameters ^60^. They found that clipped anastomoses were akin healthy normal tissue with maximal bowel wall tensile strength, whereas sutured anastomoses were significantly lower and ruptured at a lower maximal tension ^60^.

Similarly, one study also investigated the effect of time on anastomotic strength, assessing the tensile breaking strength and location using the Zwick material testing machine ^39^. Sheep trachea was anastomosed with different suturing techniques and resection lengths, and their rupture tension was tested at weekly intervals while comparing it to a normal trachea. The highest tensile strength seemed to be at four weeks postoperatively, at 273 N, with one week postoperatively being the weakest at 177 N. At six months, the tensile strength was 247 N. Overall, the stability of the trachea under load improved with time from operative intervention ^39^.

#### Anastomotic aid devices

Some studies have investigated devices that reinforce the anastomosis, with one such study using a staple line incorporated poly-ε-caprolactone (PCL) scaffold in 17 porcine bowels^43^. Three anastomoses were formed in each bowel, and a tension stretch test on postoperative day 5 revealed a maximal tensile strength of 15.7 N, which was significantly higher than the control of 12.7 N (p=0.01). The scaffold seemed to provide a protective buffer to the anastomotic site, increasing the tensile strength. However, these values are a magnitude lower than values exhibited in other studies and may need further clarity on the methodology of tension testing ^43^. Similarly, another group investigated using a soluble intraluminal prosthesis on tensile strength in porcine bowel, using clamps 5 mm apart, with positive results on durability of the anastomosis under tensile load ^40^. No conclusive studies discuss suture tension and consequent rates of leakage or complications ^41^. Studies have also compared intraluminal prostheses for bowel support during sewing and stapling (in animals) with subsequent reduction in suture tension and ischemia ^40,43^. However, leakage rates were not significantly improved with the implement, and laparoscopic techniques made it largely defunct ^62^. Additionally, recent publications on different surgical methods to reduce tension, such as Deloyers and retro-ileal techniques, have been shown to have similar (20%) anastomotic leak rates as traditional anastomotic techniques. Previous evidence had demonstrated that the Deloyers procedure had a lower leak rate (3.4-10%) than reported ^25,72^.

The existing literature does not seem to provide an objective, reproducible method to measure tension directly or indirectly in bowel anastomoses in live human subjects. Such a critical yet unaccountable variable leads to a lack of predictive accuracy for anastomotic leaks^36^. Hence, developing a reliable, clinically usable method to measure anastomotic tension accurately will provide surgeons with a tool to prevent potentially deadly complications of anastomotic leaks.

## Limitations

The bowel comprises different structural layers with different tensile properties, subject to normal anatomical variation. The serosa and subserosa play a minimal role in tensile strength, while the muscular and submucosal layers are responsible for most resistance to tension, both longitudinal and circumferential. Only a couple of studies recognize this distinction, and none investigate age, co-morbidity, and other factors that may influence the functioning and decline of these individual layers.

## Conclusion

Anastomotic leak following gastrointestinal surgery is a severe complication with devastating morbidity and mortality. Significant progress has been made in understanding and mitigating the risk factors associated with anastomotic leaks. The factors impacting anastomotic leak can be broadly categorized into patient-specific and technical factors. While recent advancements have targeted patient-related factors, much progress has not been made on the front of anastomotic tension. Tension is widely recognized for its importance in the success of bowel anastomoses but has remained subjective in its assessment. Cadaveric and animal studies that have explored maximal tensile loads are destructive in nature. A safe, reliable way to measure tension intra-operatively would be an invaluable tool in the surgical arsenal and provide a solid basis for future scientific study.

## Supporting information

PRISMA checklist

## Data Availability

All data referenced are available online in the MEDLINE Pubmed database.

## Conflict of interest

The authors declare no financial, commercial, personal, political, or academic conflicts of interest.

## Author contributions

DA and MK wrote the original draft, and conducted literature review along with AK. AK, JYW, MK, and DA helped with reviewing, editing, project administration, supervision and validation. All authors helped in conceptualization and resource management

## Acknowledgments

None

## Funding

None

## Notes

### Competing Interest Statement

The authors have declared no competing interest.

### Funding Statement

This study did not receive any funding

### Summary of Updates

Update to content, formatting, and compliance with updated PRISMA checklist. Author affiliations and list updated.

